# Wearable Sensing in Eating Episode Monitoring: An Updated Systematic Review Protocol

**DOI:** 10.1101/2024.10.11.24315295

**Authors:** Jiaying Zhou, Mingzhu Cai, Mayue Shi

## Abstract

**Introduction:** Objective and accurate dietary monitoring is critical for comprehensive dietary assessment and improving nutritional health outcomes. The rapid development of wearable sensing technology presents a promising solution for effective dietary monitoring that reduces recall bias and increases user convenience, contributing to clinical chronic disease management and nutritional research. This systematic review aims to explore the effectiveness and practicality of wearable sensors in monitoring dietary behaviours, while also examining the latest advancements in the field since 2020.

**Methods and Analysis:** Adhering to PRISMA-P guidelines, we will conduct a comprehensive search across MEDLINE, EMBASE, PubMed, IEEExplore, and Web of Science for research published between January 2020 and January 2024 Studies that involved human participants using wearable sensors for monitoring dietary intake will be included. However, studies solely discussing the development of algorithms or applications for these sensors will be excluded. Our review outcomes focused on evaluating the design of the sensors, their performance metrics, and user experience.

**Ethics and Dissemination:** Findings of this systematic review will be disseminated through peer-reviewed journals, conferences, and seminar presentations. The data used does not include individual patient data, so no ethical approval is required.

**Systematic Review Registration Number:** PROSPERO Registration ID: CRD42024531570

**Strengths and Limitations of This Study:** 1. This study will update the latest advancements in wearable sensing devices for monitoring food intake published from 2020 to date.

2. This study follows a PICOS framework to formulate research questions and inclusion/exclusion criteria.

3. This study uses a well-structured search strategy, using MeSH and equivalent terms to search across five medical and engineering databases to provide a precise, comprehensive, and multidisciplinary perspective on the topic.

4. The nature of the available data precludes the possibility of conducting a meta-analysis, limiting the ability to quantitatively synthesize study findings. However, the narrative data synthesis adheres to the Synthesis Without Meta-analysis (SWiM) guidelines, ensuring a structured approach to qualitatively integrating study outcomes.

5. The inclusion criteria were restricted to peer-reviewed journals published in English. This excludes potentially relevant conference papers and studies published in other languages, which may limit the comprehensiveness of the analysis.

## Introduction

### Description of the Condition

Dietary habits are a crucial determinant of health outcomes, significantly influencing the onset and progression of chronic diseases such as type 2 diabetes, heart disease, and obesity ^1^. Despite the clear connection between diet and health, accurately and objectively measuring food intake remains a significant challenge in nutritional science. Traditional methods such as direct observation and self-reported food diaries are not only prone to inaccuracies but also impose substantial burdens on participants, dietitians, and researchers. These limitations render traditional methods less feasible for large-scale studies ^2^. This underscores the urgent need for innovative approaches that enhance accuracy, usability, and compliance in dietary tools.

### Description of the Intervention

Recent technological advancements have significantly improved dietary monitoring, reducing respondent burden, minimizing recall bias, and enabling real-time data collection in naturalistic settings. Notable enhancements include mobile device-assisted and image-based assessments, such as food diary apps, alongside wearable sensors ^3^. Wearable sensors, in particular, are emerging as a promising tool. These devices are designed to be worn on the body and continuously monitor various aspects of dietary intake with minimal input from the user, facilitating seamless integration of dietary monitoring into everyday life ^1^.

These sensors continuously monitor behavioural signals and contextual information related to eating. For example, motion sensors detect various body movements such as hand-to-mouth movements ^4^, acoustic sensors capture chewing and swallowing sounds ^5^, and cameras can gather data on the meal context, including the type and volume of food, as well as the location and timing of the meal (by indirect video information?) ^6^.

### How the Intervention Might Work

Wearable sensors offer a seamless and objective way to collect, record, or analyse dietary data, reducing the reliance on self-reporting and thereby minimizing recall biases and inaccuracies. By providing continuous, objective data on dietary intake, these sensors not only enhance the accuracy of dietary assessments but also provide insights previously difficult to obtain ^8^. Wearable technologies have been successfully deployed in both laboratory and real-life settings. For instance, current nutritional research has already utilized a fusion of wearable sensors, combining camera, resistance and inertial sensors in a device called Automatic Ingestion Monitor v2 (AIM-2), for dietary data collection. In these studies, the AIM-2 sensor significantly reduces the labour-intensive burden of dietary monitoring while also demonstrating promising performance ^9^.

### Why it is Important to Conduct This Study

The rapid evolution of wearable technology, combined with the growing prevalence of chronic diseases, highlights the need for a comprehensive and updated review of these technologies. While insightful, previous reviews often categorize criteria based on sensors’ capabilities for automatic recording and their applicability in real-life settings^8, 10^. Consequently, these reviews overlook some innovative sensors that are still under development, potentially missing emerging technologies that could offer significant advancements.

A recent search in the Web of Science database using keywords such as "food intake" and "wearable sensors" reveals a marked increase in both research activity and publications in this field, especially notable from 2020 to 2022 as shown in Figure 1. The last systematic review in 2019, primarily focusing on motion sensors, overlooked various sensor types. Given the significant advancements in wearable sensing technology since then, an updated systematic review that encompasses a broader range of wearable sensors is essential^11^.

**Figure 1.**
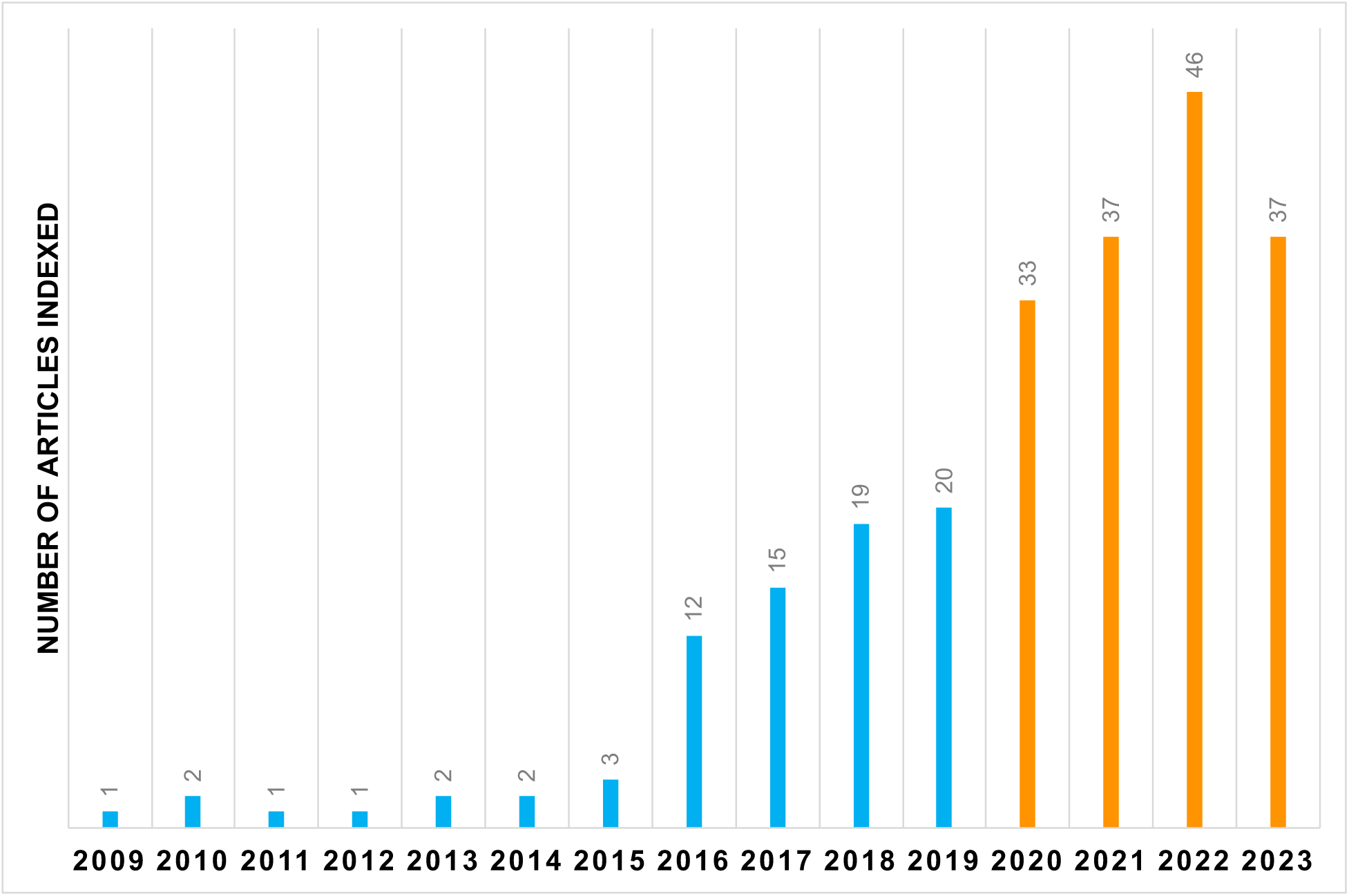
Trend in Research Publications on Wearable Sensors for Food Intake Monitoring (2009-2023). This bar graph displays the annual number of articles indexed in the Web of Science database relating to "food intake" and "wearable sensors" from 2009 to 2023. The period (2020 to 2023) planned to be included in this updated systematic review is distinctly marked with orange bars.

## Objectives

This systematic review aims to evaluate the latest advancements in wearable sensor technologies designed for monitoring dietary behaviours across various populations. The focus will be on assessing the sensor design, eating-related events detection and overall performance and applicability across different settings.

## Outcomes and Prioritization

### Primary Outcomes

This systematic review will produce detailed outcomes based on the examination of wearable sensors for monitoring food intake since 2020. The anticipated outcomes include:

1. **Sensor Technology Classification**: A comprehensive catalogue of sensor technologies identified and categorized by type and functionality within dietary monitoring.
2. **Sensor Placement Analysis**: Findings on sensor placement locations on the body, detailing the implications on the accuracy of data collected and user adherence levels.
3. **Event Detection Capability**: Results on the effectiveness of sensors in detecting eating-related events, including detailed descriptions of the types of data these sensors provide.
4. **Performance Metrics Evaluation**: Detailed evaluations of sensor performance metrics, including accuracy, precision, and sensitivity.
5. **Effectiveness in Different Settings**: Analysis of the effectiveness of these sensors in controlled lab environments versus real-life settings.

### Secondary Outcomes

The secondary outcomes will explore the broader impacts and potential advancements in wearable sensor technology for dietary monitoring:

1. **Sensor Prevalence and Dominance**: Insights into the prevalence and dominance of various sensor types throughout the review period, with a focus on emerging technologies and their impact on the market.
2. **Trend Analysis in Sensor Usage**: Comparative analysis of sensor usage trends over time, identifying shifts and developments in technology adoption since previous systematic reviews.
3. **Research Gaps and Future Directions**: A synthesis of current gaps in the research landscape, offering strategic recommendations for future technological innovations, improvements in sensor design, and data integration techniques.

## Methods and Analysis

This protocol will follow the PRISMA-P (Preferred Reporting Items for Systematic Reviews and Meta-Analysis Protocols) guidelines, which include a checklist of 17 essential items^12, 13^. These items will represent the minimum necessary components for a systematic review or meta-analysis protocol. By adhering to this standardized framework, we will ensure that our review process is both systematic and transparent.

### Eligibility Criteria for Including Studies

The eligibility criteria are framed by the PICO framework, detailing the target population, interventions, comparators, and outcomes to precisely assess the efficacy and application of wearable dietary monitoring devices in past studies.

#### Population

Inclusion Criteria:

- Individuals from all populations, including patients requiring dietary monitoring and healthy participants, irrespective of age, gender, or BMI.

Exclusion Criteria:

- Animal studies and theoretical models.

#### Intervention

Inclusion Criteria:

A ’wearable sensor’ used to monitor dietary intake specifically, refers to any apparatus worn on the body equipped with embedded sensors. These devices are designed to sense bodily or environmental conditions and record data, which can be stored directly on the device or transmitted externally via Bluetooth or other wireless communication approaches. Studies using wearable sensors designed specifically for dietary intake monitoring, which include capabilities for:

- Detecting eating events, encompassing the oral phase of digestion (bite, chewing and swallowing) and hand-to-mouth gestures.
- Providing contextual information about meals, such as food type classification and details of eating episodes. This includes time, duration, and setting of meals.
- Estimating energy intake consumed during eating events based on sensor data and algorithms.

Exclusion Criteria:

- Studies focused solely on water consumption and the pharmacological impacts of drugs.

#### Comparator

Inclusion Criteria:

- Comparisons between data collected by wearable sensors and ground-truth (reference) methods.
- Comparisons of sensor performance in lab-controlled, pseudo or actual real-life settings

#### Outcomes

Inclusion Criteria:

Wearable Sensor Performance Metrics:

- Accuracy, specificity, precision, recall, or F1-score (for Eating Detection).
- Correlation between sensor measurements and outcomes.
- Variance in estimated energy intake values. Feasibility and User Experience:
- Real-life usability.
- Intrusiveness.
- Fitting and comfortability.
- Wearing compliance. Exclusion Criteria:
- Articles without reported results from empirical research

#### Study Design and Context

Inclusion Criteria:

- Experiential studies involving human participants evaluating the use of wearable sensors in dietary monitoring, in either controlled or real-life environments.
- English-language articles in peer-reviewed journals that were published after January 1, 2020.

Exclusion Criteria:

- Review articles, commentaries and protocols, or publications before January 1, 2020.
- Studies without human involvement or focusing solely on sensor algorithms and technical details will be excluded.

### Information Sources and Search Strategy

Searches will be conducted in MEDLINE, EMBASE, PubMed, IEEExplore, and Web of Science, adhering to specific eligibility criteria. Literature search strategies using Medical Subject Headings (MeSH) and keywords related to wearable sensors and eating activities will be developed. Keywords will include ’eat’, ’food intake’, ’dietary intake’; ’monitor’, ’assess’, ’detect’, ’track’; ’wearable sensor’, ’wearable device’, ’wearable technology’, and ’smartwatch’. Please refer to Supplementary Table 1 (Appendix) for the complete search strategy for each database.

### Screen and Selection Process

The literature search results will be uploaded to Covidence^14^, a web-based tool for systematic review management. Duplicates will be automatically removed upon uploading. Our team will create and refine screening questions and forms, aligning them with our predetermined inclusion and exclusion criteria.

The selection process will start with all three authors independently reviewing titles and abstracts to check if they meet the inclusion criteria. Titles that seem to fit these criteria or whose eligibility is not clear will be selected for full report review to confirm they comply with the inclusion criteria. Conflicts on Covidence will be resolved through discussion in review team meetings, and reasons for excluding texts will be recorded. The flow diagram of the article selection process on Covidence will be shown in the PRISMA flow diagram (Figure 2).

**Figure 2.**
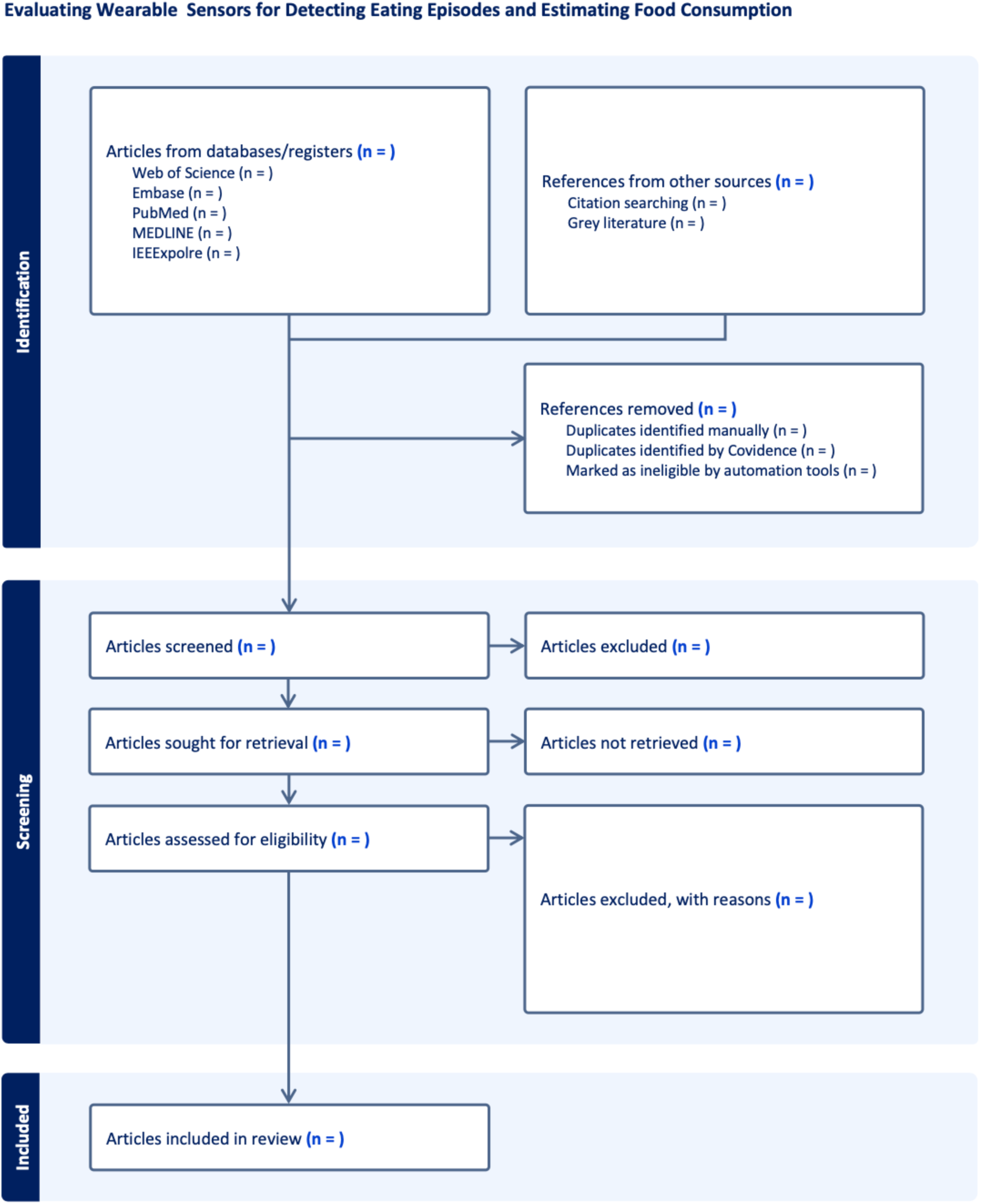
**PRISMA Flow Diagram of the Article Screening Process.**

### Data Extraction

J.Z. will spearhead independently extracting specified characteristics from selected studies into a designated Excel spreadsheet for data extraction. Following this, M.C. and M.S. will independently assess the extracted data. Any discrepancies identified during their review will be resolved in team meetings. Given the diversity of data items to be extracted and the varied focus across papers, it is expected that not all papers will contain every piece of desired information.

Pre-defined characteristics from the chosen eligible studies will be extracted into a data extraction Excel spreadsheet. The information set for extraction will include:

**Population:**

1. Population: Characterizing the demographic and health status of participants included in the study, such as age range, gender distribution, and any specific health conditions.
2. Sample Size: Specifying the number of participants enrolled and analysed in the study.

#### Intervention

3. Tested Meal: Describing the types of food or diet consumed by the participants for sensor data collection.
4. Study Period: Indicating the duration over which the study was conducted.
5. Sensor Types: Identifying the specific sensor(s) employed within wearable devices for eating detection.
6. Number of Sensors: Specifying the quantity of sensors used in the study.
7. Sensor Design: Describing the engineering and aesthetic aspects of the sensor, including its wearability, size, and discreetness.
8. Sensor Placement: Noting the body position of sensor placement where the wearable sensor(s) are positioned, such as wrist, neck, or ear.
9. Eating Event Detection: Detailing the sensor-detected eating activities, including the types of eating motions captured.
10. Meal Contextual information: Detailing the contextual information of the meal that can be detected or provided by the sensor data, such as time of day, duration of meal, or setting.
11. Level of Automation: Describing if the wearable sensor in food intake monitoring and data analysis is fully automatic, semi-automatic or manual-dependent.
12. Study Setting: Classifying if the study is set up in a lab-controlled, pseudo-, or actual free-living setting.

#### Comparisons

13. Ground-Truth Method(s): Describing the benchmark method(s) used for assessing sensor performance and making comparisons.
14. Real-World Usability: Comparing the data collected by the wearable sensor in free-living and lab-controlled environments, assessing adaptability and accuracy in different settings.

#### Outcomes

15. Performance Metric(s): Detailing the efficacy of wearable sensors in recognizing eating behaviours, including precision, accuracy, recall, and any other relevant metrics, provided they are clearly stated as text, within tables, or illustrated in figures. It is important to note that our review reported only the metrics provided by the original studies, without performing any new calculations.
16. User Experience: Describing the participants’ feedback on the wearable sensor, including comfort, ease of use, wearing compliance and overall satisfaction, as well as any impact on their normal eating behaviour.

### Quality Control

#### Assessment of Risk of Bias in Included Studies

In this systematic review, a comprehensive assessment of the risk of bias in individual studies will be conducted to ensure the reliability and validity of our findings. We will follow the structured guidelines provided in Chapter 25 of the Cochrane Handbook for Systematic Reviews of Interventions, utilizing specific tools tailored to the type of studies under review ^15^:

- **ROBINS-I tool** for non-randomized studies, evaluating domains such as confounding, selection of participants, classification of interventions, deviations from intended interventions, missing data, measurement of outcomes, and selection of reported results ^16^.
- **RoB 2 tool** for randomized studies, examining domains including the randomization process, deviations from intended interventions, missing outcome data, measurement of the outcome, and selection of the reported result ^17^.

The risk of bias assessment will be carried out at both the study and outcome levels to identify biases that could impact the overall study or specific outcomes. The results from these assessments will be crucial in the data synthesis process:

- **Categorization of Studies:** Each study will be categorized based on its assessed risk of bias—low, moderate, high, or critical. This categorization will directly influence the weight that each study contributes to the synthesis.
- **Interpretation of Findings:** The categorization will also play a vital role in interpreting the review’s findings, especially when integrating results from studies with high or critical risk of bias.

#### Visualization of Bias Assessment

The results of the risk of bias assessments will be visualized using robvis, a tool that generates high-quality, publication-ready figures summarizing these assessments for systematic reviews ^18^. This visualization tool allows for customization according to the specific assessment tool used, such as RoB 2 or ROBINS-I, ensuring that our review’s risk of bias visualizations are both informative and tailored to our specific methodology. These visualizations will aid in transparent reporting and a detailed discussion of the biases inherent in the included studies.

### Data Synthesis

Since quantitative synthesis is not appropriate in this systematic review, a narrative synthesis will be systematically conducted according to the Synthesis Without Meta-analysis (SWiM) guidelines, presenting information in both text and tabular formats to summarize and elucidate the characteristics and outcomes of the studies included ^19^.

1. **Grouping Studies for Synthesis:** Studies will be grouped by wearable sensor types, sensing technology, sensor placements, eating event detection methods, and setting (lab-controlled vs. real-life). This approach highlights trends in wearable sensor development for food intake monitoring, focusing on sensor placement and application effectiveness.
2. **Standardised Metrics and Transformation:** Performance metrics from each study will be extracted as reported, highlighting the most effective algorithms or methods as identified by the studies or by J.Z. based on the highest metric value. Our review will relay the original study metrics without new calculations.
3. **Investigation of Heterogeneity:** Heterogeneity will be explored through informal methods, such as organizing tables or figures by study design, subpopulations, intervention components, and setting factors. This approach is indicative and used to identify potential sources of variability among study findings, recognizing the exploratory nature of these methods.
4. **Reporting Results:** Results will be systematically organized and presented in tables, outlining the design and major findings of each study. These will be complemented by visual aids, including bar and pie charts, to effectively illustrate key data. For instance, the charts will showcase the prevalence of different types of wearable sensors and their placement on the body, providing a clear and engaging overview of the latest trends and applications in dietary monitoring.
5. **Comparisons to Previous Reviews:** This updated systematic review will conduct an in-depth comparison of the latest findings in wearable sensor technologies for dietary monitoring with those from previous reviews, to identify advancements and trends. It will analyse trends in wearable sensor design, spotlighting dominant technologies and significant new entrants, and providing a comprehensive overview of the technology landscape in this field. The review will also focus on comparing the highest performance metrics—such as accuracy, sensitivity, and precision—from current studies against those documented in earlier reviews. Moreover, it will identify areas where further research could enhance sensor utility for dietary monitoring. Based on the identified gaps, the review will propose future research directions. These will focus on potential enhancements in sensor design and the exploration of new areas for technological innovation in dietary monitoring.

### Confidence in cumulative evidence

The strength of the evidence will be assessed using the GRADE approach. It will evaluate the quality of evidence across studies and the strength of recommendations regarding the use of wearable sensors for monitoring food intake. This framework considers:

1. Risk of Bias: Assesses the potential for bias within individual studies.
2. Inconsistency: Examines the variability in effect estimates across studies.
3. Indirectness: Evaluate the relevance of the evidence to the research question and the population of interest.
4. Imprecision: Considers the confidence in effect estimates based on confidence intervals and sample sizes.
5. Publication Bias: Assesses the likelihood of selective publication of studies.

Given that a quantitative synthesis (meta-analysis) is not appropriate, a narrative synthesis will be conducted. This will involve summarizing findings from individual studies in text and tabular formats, highlighting the key characteristics and outcomes. The evidence for each primary and secondary outcome will be summarized and categorized into high, moderate, low, or very low confidence using the GRADE criteria. This assessment will guide our conclusions and recommendations regarding the use of wearable sensors in dietary monitoring.

## Ethics and dissemination

This systematic review does not necessitate ethical approval, as it relies on already published studies containing non-identifiable data. The outcomes of the review will be shared through publication in a peer-reviewed journal and presentations at conferences and seminars.

## Discussion

The previous systematic review, published five years ago, was pioneering in summarizing the application of upper limb motion tracking sensors—either as standalone or multi-sensor devices—for the objective assessment of eating behaviour ^11^. Since then, the field has seen substantial growth. In the past four years, a search on the Web of Science database alone using the terms "food intake" and "wearable sensors" has yielded 153 new publications. Given this significant advancement in research, an update to the review is essential to capture the latest developments and any new emerging technology in the evolving landscape of wearable technology in dietary monitoring. The results of this systematic review will contribute to future research into the development of wearable-based methods of tracking eating and to improve the management of diet-related diseases in the future.

## Data Availability

N/A

## Declarations

### Patient Consent for Publication

Not required.

### Author’s Contributions

M.C. and M.S. designed and directed the project; M.C. and M.S. acquired funding as Co-PIs; J.Z., M.C. and M.S. developed and wrote this systematic review protocol.

## Appendix

**Supplementary Table 1.** Database-Specific Search Queries.

|  |  |
| --- | --- |
| MEDLINE | ((("eat" or "food intake" or "dietary intake" or "diet" or "ingest" or "feed") And ("monitor" or "assess" or "detect" OR "track") And ("wearable" or "wearable device" or "wearable sensor" or "smart watch" or "smartwatch" or "wearable technology" OR "fitness tracker"))) |
| EMBASE | ((("eat" or "food intake" or "dietary intake" or "diet" or "ingest" or "feed") And ("monitor" or "assess" or "detect" OR "track") And ("wearable" or "wearable device" or "wearable sensor" or "smart watch" or "smartwatch" or "wearable technology" OR "fitness tracker"))) |
| PubMed | ((("Nutrition Assessment"[Mesh] OR "Nutrition Surveys"[Mesh] OR "Diet, Food, and Nutrition"[Mesh] OR "Eating"[Mesh] OR "Feeding Behavior"[Mesh] OR "Food Intake" OR "dietary behaviour" OR "dietary intake" OR "ingest" OR "eating" OR "feed") AND ("monitor" or "assess" or "detect" OR "track") AND ("Wearable Electronic Devices"[mesh] OR "smart watch" OR "smartwatch" OR "wearable sensor" OR "wearable device" or "wearable technology" OR "fitness tracker"))) |
| IEEEExplorer | ((("feed" OR "eat*" OR "diet*" OR "food" OR "ingest*" OR "food intake" OR "dietary intake") AND ("monitor*" OR "assess*" OR "detect*" OR "track*")) AND ("wearable" OR "wearable device" OR "wearable sensor" OR "smart watch" OR "smartwatch" or "wearable technology" OR "fitness tracker"))) |
| Web of Science | ((((PY=(2020-2024)) AND TS=((( "eat*" OR "diet*" OR "food" OR "ingest*" OR "food intake" OR "dietary intake") AND ("monitor*" OR "assess*" OR "detect*" OR "track*")) AND ("wearable device" OR "wearable sensor" OR "smart watch" OR "smartwatch" or "wearable technology" OR "fitness tracker"))))) AND LA=(English)) AND DT=(Article) |

## Reference

1 Bell BM, Alam R, Alshurafa N, et al. Automatic, wearable-based, in-field eating detection approaches for public health research: a scoping review. NPJ digital medicine 2020;3:38 doi:10.1038/s41746-020-0246-2.

2 Naska A, Lagiou A, Lagiou P. Dietary assessment methods in epidemiological research: current state of the art and future prospects [version 1; peer review: 3 approved]. F1000 research 2017;6:926 doi:10.12688/f1000research.10703.1.

3 Burrows TL, Rollo ME. Advancement in Dietary Assessment and Self-Monitoring Using Technology. Nutrients 2019;11:1648 doi:10.3390/nu11071648.

4 Chun KS, Jeong H, Adaimi R, et al. Eating Episode Detection with Jawbone-Mounted Inertial Sensing. EMBC Jul 1, 2020;2020:4361–4 doi:10.1109/EMBC44109.2020.9175949.

5 Khan MI, Acharya B, Chaurasiya RK. iHearken: Chewing sound signal analysis based food intake recognition system using Bi-LSTM softmax network. Computer Methods and Programs in Biomedicine 2022;221 doi:10.1016/j.cmpb.2022.106843.

6 Alshurafa N, Zhang S, Romano C, et al. Association of number of bites and eating speed with energy intake: Wearable technology results under free-living conditions. Appetite 2021;167 doi:10.1016/j.appet.2021.105653.

7 Vu T, Lin F, Alshurafa N, et al. Wearable Food Intake Monitoring Technologies: A Comprehensive Review. Computers (Basel) 2017;6:4 doi:10.3390/computers6010004.

8 Hiraguchi H, Perone P, Toet A, et al. Technology to Automatically Record Eating Behavior in Real Life: A Systematic Review. Sensors (Basel, Switzerland) 2023;23:7757 doi:10.3390/s23187757.

9 Doulah A, Ghosh T, Hossain D, et al. “Automatic Ingestion Monitor Version 2” – A Novel Wearable Device for Automatic Food Intake Detection and Passive Capture of Food Images. IEEE J Biomed Health Inform 2021;25 doi:10.1109/jbhi.2020.2995473.

10 Bell BM, Alam R, Alshurafa N, et al. Automatic, wearable-based, in-field eating detection approaches for public health research: a scoping review. NPJ digital medicine 2020;3:38 doi:10.1038/s41746-020-0246-2.

11 Heydarian H, Adam M, Burrows T, et al. Assessing Eating Behaviour Using Upper Limb Mounted Motion Sensors: A Systematic Review. Nutrients 2019;11:1168 doi:10.3390/nu11051168.

12 Shamseer L, Moher D, Clarke M, et al. Preferred reporting items for systematic review and meta-analysis protocols (PRISMA-P) 2015: elaboration and explanation. BMJ 2015;349:7647 doi:10.1136/bmj.g7647.

13 Moher D, Shamseer L, Clarke M, et al. Preferred reporting items for systematic review and meta-analysis protocols (PRISMA-P) 2015 statement. Systematic reviews 2015;4 doi:10.1186/2046-4053-4-1.

14. Monique Grenier. Covidence 2024.

15 Higgins JPT, Thomas J, Chandler J, et al. Cochrane Handbook for Systematic Reviews of Interventions. Newark: John Wiley & Sons, Ltd 2019.

16 Sterne JA, Hernán MA, Reeves BC, et al. ROBINS-I: a tool for assessing risk of bias in non-randomised studies of interventions. BMJ (Online) 2016;355:i4919 doi:10.1136/bmj.i4919.

17 Sterne JAC, Savović J, Page MJ, et al. RoB 2: a revised tool for assessing risk of bias in randomised trials. BMJ (Online) 2019;366:l4898 doi:10.1136/bmj.l4898.

18 McGuinness LA, Higgins JPT. Risk-of-bias VISualization (robvis): An R package and Shiny web app for visualizing risk-of-bias assessments. Research Synthesis Methods 2021;12:55–61 doi:10.1002/jrsm.1411.

19 Campbell M, McKenzie JE, Sowden A, et al. Synthesis without meta-analysis (SWiM) in systematic reviews: reporting guideline. BMJ 2020;368:l6890- doi:10.1136/bmj.l6890.

